# Navigating Health Insurance Selection for in Vitro Fertilization (IVF) Benefits: A Study Protocol

**DOI:** 10.1101/2025.03.27.25324807

**Authors:** Olumayowa Dayo, Victoria Turcotte, Breanna Reyes, Ricardo E. Flores Ortega, Bonnie N. Kaiser, Gregory A. Aarons, Sara B McMenamin, H. Irene Su, Sally A. D. Romero

**Author notes:** **Corresponding author:** Author Name: Sally A. D. Romero, PhD, MPH Academic Affiliation: Department of Obstetrics, Gynecology and Reproductive Sciences, University of California San Diego Mailing Address: 3855 Health Sciences Dr MC 0901, La Jolla, CA 92093.

## Abstract

**Introduction:** A large public university added health insurance coverage of 50% co-insurance for up to two cycles of in vitro fertilization (IVF) to eligible faculty and staff.

**Methods:** We describe the design and conduct of a randomized controlled trial to evaluate the effectiveness of a health insurance educational intervention on health insurance literacy and IVF benefit utilization. The intervention materials included 1) Key insurance terms; 2) Examples of premiums and deductibles across the insurance plan options; 3) Examples of how premiums and deductibles affect out-of-pocket costs; and 4) A guide to find in-network providers/facilities. The primary outcome is health insurance literacy. Secondary outcomes are IVF services and insurance benefit utilization, out-of-pocket costs, and financial hardship related to fertility care. We collected validated patient-reported outcomes at three timepoints over 1 year. We will integrate mixed methods data to explore whether the intervention was effective, feasible, acceptable, and appropriate.

**Results:** Among 394 faculty and staff screened, 217 (55%) reproductive-aged (18 to 50 years) employees consented, completed the baseline survey and were randomized in a 2:1 fashion. Participants were female (81%), married (63%), and worked as a staff employee (72%). Approximately 39% reported an infertility diagnosis, and 28% had undergone prior IVF treatment. Participants reported feeling slightly confident when using their health insurance plans and moderately confident being proactive when using their health insurance plans.

**Discussion:** Our goal is to improve health insurance literacy and utilization of health insurance benefits for IVF care, thereby expanding family-building options for reproductive-aged individuals.

## 1. Introduction

Infertility is a disease of the reproductive system, characterized by the inability to achieve a successful pregnancy and/or the need for medical intervention to achieve a successful pregnancy either as an individual or with a partner [1]. Common reasons for fertility care include polycystic ovary syndrome, primary ovarian insufficiency, uterine fibroids, male factor infertility, same-sex individuals, single parents and unexplained infertility. Data from the National Survey of Family Growth (2015-2019) highlight that infertility affects approximately 14.4% of women (aged 15 to 49) and 11.4% of men (aged 15 to 49) in the United States [2]. Infertility has been linked to higher morbidity and mortality [3, 4].

Evidence-based fertility treatments [i.e., medications, surgical procedures, intrauterine insemination (IUI), and the most successful assisted reproductive technology (ART), which includes in vitro fertilization (IVF)] effectively decrease infertility and improve family building and quality of life [5, 6]. Yet, research has shown that only one-quarter of the need for ART in the U.S. is being met.[7] IVF treatment costs are high ($15,000 to $30,000 on average) and typically not covered by health insurance. Treatment costs are a major barrier to infertility care and contribute to financial burden, distress and delayed treatment and low access to care [7–10].

Currently, insurance coverage for fertility treatments varies by state, and most do not require private insurance or federal health insurance (Medicare, Medicaid) to cover fertility treatments [11]. Therefore, the inclusion of health insurance benefits for infertility care can be considered a value-add for employees with private insurance plans. Even with the increase in insurance coverage for fertility care, navigating health insurance benefits can be complex, limiting access to available treatments [12]. Understanding the complex network of health insurance, specifically fertility treatment coverage, is essential to provide comprehensive health plans for newly established benefits.

Health insurance literacy refers to a person’s ability to seek, obtain, and comprehend health insurance plans, and once enrolled, to effectively use their insurance to seek appropriate health care services [13]. Research has shown that approximately half of adults consider themselves to have poor health insurance literacy and lack confidence in their ability to use their insurance [14]. Moreover, even among insured adults, lower health insurance literacy was associated with delayed or foregoing of care due to cost [15] and higher medical debt [16]. Despite data linking health insurance literacy with health care utilization [17], few interventions have been developed to improve health insurance literacy. Results from two recently completed pilot clinical trials among young cancer survivors found that a patient navigator delivered psychoeducation on health insurance policy and coverage was feasible, acceptable and improved health insurance literacy [18, 19]. To date, no health insurance literacy interventions related to IVF care have been conducted.

During health insurance plan open enrollment, a public research university added coverage for an IVF benefit (50% co-insurance for up to two cycles of IVF) for all five health plans offered to eligible faculty and staff starting in the 2023 benefit year. The insurance benefit was intended to be equal across all plans, yet due to plan-specific differences in cost-sharing (e.g., monthly premiums, deductibles), prior authorization processes, and in-network providers and facilities for IVF care, how employees access and utilize these new IVF benefits may vary by plan. Navigating these differences to make an informed choice may also be challenging due to limitations in health insurance literacy. Hence, the purpose of this study was to evaluate the impact of an IVF health insurance benefit educational intervention on health insurance literacy and utilization of IVF insurance benefits. We hypothesize that the intervention will be more acceptable and satisfactory to employees than university provided standard materials, and result in increased health insurance literacy and utilization of IVF insurance benefits.

## 2. Methods

### 2.1 Study Design

A randomized controlled trial was conducted to evaluate the impact of a health insurance educational intervention on health insurance literacy and IVF benefit utilization while observing and gathering qualitative data on implementation guided by the policy-optimized exploration, preparation, implementation, sustainment (EPIS) framework [20]. The institutional review board (IRB) at the University of California, San Diego approved the study.

The trial was conducted over 13 months at a single public research university. Enrollment began in November 2022, and study participant assessments were completed by December 2023. The target accrual goal was 200 participants. Participants completed a 30-minute baseline questionnaire prior to randomization and at follow up 1, they reported their health plan selection and satisfaction with their decision. Follow up 2 was completed 7 to 12 months after health plan enrollment and assessed fertility history, financial well-being, health insurance literacy, and IVF utilization. Semi-structured interviews via telephone, in person or Zoom were conducted in a subset of participants about experiences with the intervention, health plan selection and accessing and utilizing IVF services. The intervention was designed to be an educational guide to navigate choosing one of five health plans with IVF coverage. During the open enrollment period, participants randomized to the intervention condition were emailed the health insurance education intervention, and those randomized to the usual care condition were emailed the usual care intervention.

### 2.2 Educational Intervention

The intervention consisted of a 12-page PDF health insurance educational guide presented in English and Spanish and highlighting key health insurance literacy terms and considerations when choosing among the university’s five health plan options for IVF coverage (See Appendix). The intervention was grounded in the Integrated Behavioral Model [21], which takes a behavioral economics perspective, and Anderson’s Behavioral Model for Health Services Use [22] by targeting individual cognition and self-efficacy predisposing factors and socio-environmental enabling factors to improve health insurance literacy and promote intentions and utilization of IVF services.

The four content areas covered in the intervention were:

1. Definition of key insurance terms: Definitions were retrieved from the glossary of health coverage and medical terms from healthcare.gov for the following seven health coverage terms: allowed amount, co-insurance, co-payment, deductible, in-network, out-of-pocket maximum, and premium.
2. Differences in premiums, deductibles, and eligibility criteria for the IVF benefit (based on insurer’s definition of infertility) across the five 2023 offered health plans. Since premiums and deductibles vary by salary categories, materials were tailored to salary category [(1) $65,000 and below; (2) $65,001-129,000; (3) $129,001-194,000; and (4) $194,001 and above.]
3. Comparison of total out-of-pocket costs for the member across the five plans. We developed a graphic summary of how premiums, deductibles, and IVF services costs at 50% co-insurance factor into the out-of-pocket costs for IVF for the member.
4. Guide to finding in-network providers and facilities for IVF care via four steps: (a) decide on a medical group for HMO subscribers, (b) call health plan or searching online to identify which reproductive endocrinologists are in-network, (c) call health plan to ask which IVF facilities are in network and providing a CPT code for IVF services, and (d) ask about the process to submit claims for reimbursement if out-of-network providers or facilities are used.

The intervention arm received the educational intervention and standard open enrollment guide. The usual care arm received a one-page open enrollment guide from the university describing the expanded fertility benefit and providing links to 1) the university benefits open enrollment website; 2) virtual benefits advisor chatbot; and 3) a virtual benefits office hours during the open enrollment period.

### 2.3 Inclusion and Exclusion Criteria

Participants were eligible for the study if they were university faculty or staff eligible for health insurance coverage by the institution, English or Spanish speaking, reproductive-aged adults (18 to 50 years), and had the intention to grow their family in the next year.

### 2.4 Procedures

University faculty and staff emails were identified through the public university website ‘Search Faculty/Staff’ function. During open enrollment for the 2023 benefit year, study investigators e- mailed university faculty and staff to explain the newly enhanced IVF insurance benefit, the study, and providing a link for interested participants to complete an online eligibility screening. Eligible faculty and staff were taken directly to an online consent form and baseline questionnaire; those who were ineligible received a message stating their ineligibility and a thank you message. After completion of the baseline questionnaire, participants were randomized in a 2:1 fashion. Those randomized to the intervention were emailed the health insurance education guide and the standard open enrollment material while those randomized to the usual care condition were emailed the standard university-provided open enrollment materials. All participants were emailed follow-up questionnaires at two time points: 1 month and 7 months post-enrollment. Among participants who completed the follow-up 2 questionnaire, we purposefully sampled a subset to complete semi-structured interviews via telephone, in person or Zoom. Study questionnaire data were collected and managed using REDCap (Research Electronic Data Capture) electronic data capture tools hosted at University of California, San Diego.[23, 24]

### 2.5 Randomization

We randomized participants to the intervention versus usual care condition in a 2:1 fashion using a random number generator.

### 2.6 Primary Outcome

The primary outcome is the improvement of health insurance literacy at follow up 2, as measured by 9 items from the Health Insurance Literacy Measure that assess two constructs of health insurance literacy: 1) confidence using and 2) being proactive when using health insurance plans [25] at baseline and follow up 2. The measure includes questions, such as “how confident are you that you know what to do if your health plan refuses to pay for a service that you think should be covered”, “when using your health insurance plan, how likely are you to look to member services to tell you what services your health plan covers”. Items will be scored from 0 (not at all confident) to 3 (very confident). Higher scores indicate better health insurance literacy.

### 2.7 Secondary Outcomes

At baseline, we will collect self-reported information on health plan selection, factors influencing health plan selection, reproductive health history, pregnancies and pregnancy intentions, and fertility assessments and treatments. We will also collect demographic information (e.g., race/ethnicity, sexual orientation, marital status, education, employment status, type of university employee, and household income).

At follow up 1, we will collect health plan selection and reasons why the specific health plan was selected for 2023. We will also collect information on the resources participants may have contacted during Open Enrollment regarding information about the IVF benefit and time spent with these resources and satisfaction with these resources. We will also assess participants’ likelihood to compare health insurance plans at the time of Open Enrollment as measured by 6 items from the Health Insurance Literacy Measure [25]. This measure includes questions, such as “When comparing health plans how likely are you to: Find out if the plans cover unexpected costs such as hospital stays”; Understand what you would have to pay for emergency department visits?” Items will be scored from 0 (not at all confident) to 3 (very confident). Higher scores indicate better health insurance literacy. We will also assess participants’ confidence with understanding basic health insurance terms (i.e., allowed amount, co-insurance, co-payment, deductible, in-network, out-of-pocket maximum, and premium) with response options of “not at all confident” to “very confident”. Additionally, acceptability, feasibility, appropriateness, and satisfaction with the educational materials will be measured using the Intervention Appropriateness Measure, Acceptability of Intervention Measure, and Feasibility of Intervention Measure [26]. These measures have substantive and discriminant content validity and high test-retest reliability [26].

At follow up 2, we will assess participants’ confidence with understanding basic health insurance terms (i.e., allowed amount, co-insurance, co-payment, deductible, in-network, out-of-pocket maximum, and premium) with response options of “not at all confident” to “very confident”. Additionally, we will collect self-report of utilization of IVF services in 2023, utilization of health insurance for IVF services and out-of-pocket expenses related to IVF services in 2023 and prior to 2023. Medical financial hardship related to the cost of IVF services in 2023 will be measured using an adapted version of the 15-item Economic Strain and Resilience in Cancer tool [27]. Medical financial hardship items will be scored from 0 (least hardship) to 10 (most severe hardship). Higher scores indicate worse financial hardship. Additional medical financial hardship questions from the Childhood Cancer Survivorship Study Health Insurance Survey (10 items for cost worry, 9 items for medical expenses, and 8 items for worry) [28] and from the National Health Interview Survey (2 items for material hardship, 2 items for psychological hardship and 3 items for behavioral hardship) [29] were modified to be related to IVF services received in 2023. Open-ended questions ascertained participants’ experiences with learning about or using the IVF insurance benefit as well as any tips they would give other employees related to learning about or using the IVF insurance benefit.

Qualitative semi-structured interviews used open-ended questions to assess the impact of the educational intervention (e.g., likes, dislikes, satisfaction with information, need for additional information/resources, etc.) (6 items) and participant experiences with selecting a health plan after open enrollment (3 items), IVF benefit utilization, IVF services and related out-of-pocket costs (8 items).

### 2.8 Analytic Approach

Quantitative data: The primary analysis will be intention-to-treat. We will summarize continuous variables as medians and means, as appropriate, and categorical variables as proportions. Using Student’s t-test, Wilcoxon rank sum test, Chi-square test of proportions and Fisher’s Exact tests, as appropriate, we will compare outcomes by intervention condition. We will estimate effect sizes using unadjusted logistic and linear regression models.

Qualitative data: Interview recordings will be transcribed. We will conduct thematic analysis of qualitative data (open-ended survey responses, interviews) in MaxQDA software [30]. We will identify inductive themes (arising from the data) anticipated to be related to health insurance literacy, IVF benefit access and utilization and deductive themes (arising from literature) on implementation of the IVF benefit, guided by the EPIS framework: innovation (insurance benefit characteristics and fit), inner context (employer, clinics, providers), outer context (employer, insurers, employees), and factors that bridge these levels (contracts between insurer and clinics/facilities).

Integration of qualitative and quantitative data: Following the taxonomy of mixed methods designs [31], the structure of these data is qual. + quant. Quantitative data complement qualitative findings. The data are combined at the interpretive level, while each data set remains analytically separate [32]. Triangulation of these data aims to (1) explain the process through which participants who may use IVF benefits select a health plan and access the benefits; (2) describe the effectiveness of the intervention; and (3) identify future adaptations of the intervention.

To address missing data, we will perform sensitivity analyses and apply robust data analysis strategies to evaluate data losses. We will first explore whether missingness is associated with observed variables (e.g. randomization arm and the baseline characteristics) by comparing participants with complete and incomplete data. For outcome variables with missing data, we will impute the missing outcome data using multiple imputation methods.

### 2.9 Sample Size

Sample size was based on feasibility in this pilot trial. We aimed to enroll and randomize 200 participants into the trial. For the semi-structured interviews, the target accrual goal was 40 participants or until saturation is attained. Saturation (the point at which additional data collection does not yield new insights) is the gold standard for sampling [33].

## 3. Results

We invited 19,136 university faculty and staff to participate via email. Of these, 394 interested faculty and staff completed the eligibility screener, and 131 were deemed ineligible due to age, family building intentions or employer-based health insurance plan not inclusive of the new IVF benefit. Two hundred and seventeen eligible faculty and staff completed the informed consent and baseline questionnaire, with 142 randomized to the intervention arm and 75 to the usual care arm (See Figure 1). Overall, study participants primarily identified as female (81%) and heterosexual (87%), were married (63%), had a post-graduate college education (66%) and worked as a staff employee (72%). Eighteen percent of participants identified as having Spanish/Hispanic/Latinx heritage, and 45% identified as white, 29% as Asian and 26% of other races. At baseline, 39% reported an infertility diagnosis, 60% had previously seen a fertility specialist, and 28% had undergone prior IVF treatment. On average, participants reported feeling slightly confident using their health insurance plans [mean (standard deviation) 1.03 (0.76)] and moderately confident being proactive when using their health insurance plans [2.03 (0.67)]. Baseline characteristics did not vary by group (p>0.05) (Table 1).

**Figure 1.**
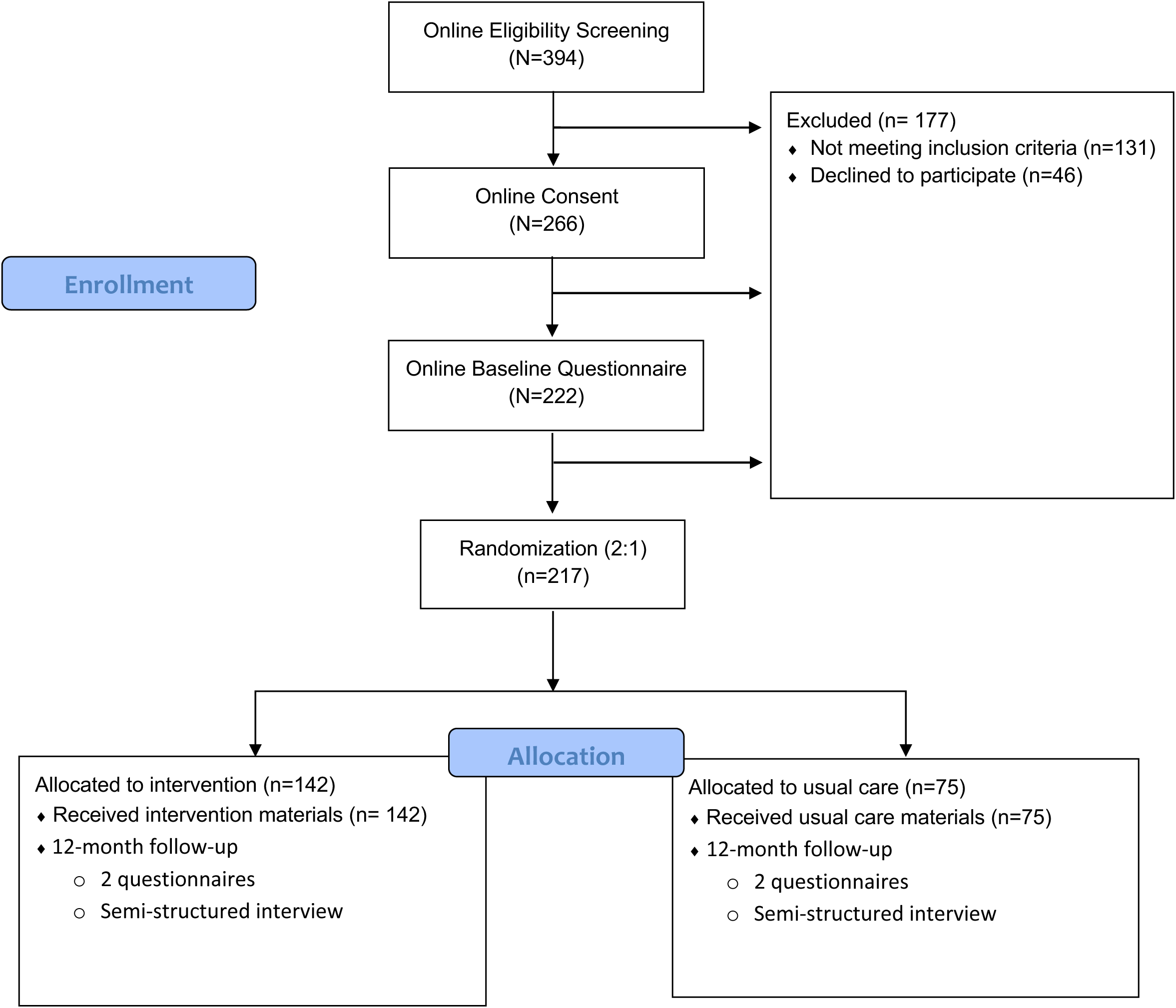
Consort diagram

**Table 1.** Participant Baseline Characteristics.

|  | Total<br>N=217<br>N (%) | Intervention<br>N=142<br>N (%) | Usual Care<br>N=75<br>N (%) | P-value |
| --- | --- | --- | --- | --- |
| **Demographics** |  |  |  |  |
| Race |  |  |  | 0.63 |
| White | 98 (45.2) | 61 (43.0) | 37 (49.3) |  |
| Asian | 63 (29.0) | 42 (29.6) | 21 (28.0) |  |
| Other | 56 (25.8) | 39 (27.4) | 17 (22.7) |  |
| Hispanic/Latino Ethnicity | 39 (18.0) | 27 (19.0) | 12 (16.0) | 0.58 |
| Education |  |  |  | 0.90 |
| Graduated from college, some college<br>or from high school | 75 (34.1) | 48 (33.8) | 26 (34.7) |  |
| Postgraduate school or degree | 142 (65.9) | 94 (66.2) | 49 (65.3) |  |
| Employee Type |  |  |  | 0.53 |
| Staff | 159 (73.3) | 106 (74.6) | 53 (70.7) |  |
| Faculty | 58 (26.7) | 36 (25.4) | 22 (29.3) |  |
| Household Income |  |  |  | 0.30 |
| Less than \$129K | 81 (37.3) | 57 (40.1) | 24 (32.0) | |
| \$129,001 - \$194,000 | 42 (19.4) | 28 (19.7) | 14 (18.7) | |
| More than \$194,000 | 48 (22.1) | 26 (18.3) | 22 (29.3) | |
| Not disclosed | 46 (21.2) | 31 (21.8) | 15 (20.0) |  |
| Sex at birth |  |  |  | 0.86 |
| Female | 176 (81.1) | 116 (81.7) | 60 (80.0) |  |
| Male | 41 (18.9) | 26 (18.3) | 15 (20.0) |  |
| Sexual Orientation |  |  |  | 0.89 |
| Heterosexual/Straight | 189 (87.1) | 124 (87.3) | 65 (86.7) |  |
| LGBTQ/other | 28 (12.9) | 18 (12.7) | 10 (13.3) |  |
| Marital Status |  |  |  | 0.67 |
| Married | 137 (63.1) | 89 (62.7) | 48 (64.0) |  |
| Partnered | 36 (16.6) | 22 (15.5) | 14 (18.7) |  |
| Single | 44 (20.3) | 31 (21.8) | 13 (17.3) |  |
| **Fertility History** |  |  |  |  |
| Infertility diagnosis | 85 (39.2) | 52 (36.6) | 33 (44.0) | 0.31 |
| Fertility specialist visit | 130 (59.9) | 84 (59.7) | 46 (61.9) | 0.77 |
| IVF with frozen embryo transfer | 61 (28.1) | 40 (28.5) | 21 (28.3) | 1.00 |
| Ovulation induction | 70 (32.3) | 47 (33.4) | 23 (31.0) | 0.76 |
| Fertility preservation embryo/oocyte | 43 (19.8) | 28 (19.9) | 15 (20.2) | 1.00 |
| Donor egg/surrogacy | 8 (3.7) | 5 (3.6) | 3 (4.0) | 1.00 |
| Surgery | 21 (9.7) | 13 (9.2) | 8 (10.8) | 0.81 |
| **Health Insurance Literacy\*** |  |  |  |  |
| Confidence using health insurance plans,<br>*mean (SD)* | 1.03 (0.76) | 1.06 (0.73) | 0.98 (0.81) | 0.49 |
| Being proactive when using health<br>insurance plans, *mean (SD)* | 2.03 (0.67) | 2.02 (0.69) | 2.05 (0.65) | 0.81 |

\*Health insurance literacy scores ranged from 0 “not at all confident” to 3 “very confident”

## 4. Discussion

Although most insurance plans cover some procedures related to infertility treatments, many plans do not include IVF treatments, making it financially inaccessible to many individuals [11]. Insurance benefits are complex and challenging to navigate, and health insurance utilization for needed services can be influenced by an individual’s health insurance literacy [14, 15, 17]. Through conducting a randomized controlled trial, we hypothesize that the intervention will be acceptable and satisfactory and will result in increased health insurance literacy and utilization of IVF insurance benefits and services.

While this clinical trial is key to understanding the effects of navigating a newly introduced IVF insurance benefit, there are some limitations. This study focused on a sample population of university staff and faculty from one of the twelve campuses impacted by this new employee enhanced insurance benefit. Participants were recruited by mass emailing potentially interested faculty and staff, which may result in a sample of reproductive-aged employees seeking IVF services and enthusiastic about gaining health insurance literacy knowledge, and this may not be generalizable to other university employees at other campuses.

Despite these limitations, we expect to find that individuals in the intervention arm will experience an increase in health insurance literacy which, in turn, may lead to higher utilization of IVF insurance benefits and services. This study will help us learn if the intervention is effective, what adaptations to the intervention may be needed to improve effectiveness, and what facilitators can help the intervention become standardized material for open enrollment periods. If successful, this pilot trial may allow for further evaluation of a larger employee population to analyze the impact of the new IVF benefit across all campuses.

This study has the potential to improve reproductive health insurance policies and accessibility of an enhanced IVF benefit provided through employer-based or private insurance plans. Information on IVF utilization and health insurance literacy may be adapted and incorporated into standard benefit enrollment platforms and impact the understanding and utilization of IVF insurance benefits.

## Data Availability

All data produced in the present study are available upon reasonable request to the authors.

## Acknowledgements

The authors would like to thank the study participants and research staff for their contributions to this study.

## Funding and Research Support

Research reported in this article was supported through funding by the National Institutes of Health / National Center for Research Resources (UL1TR001442) and National Research Service Award (T32 HD007203). The statements presented in this work are solely the responsibility of the authors and do not necessarily represent the views of the National Institutes of Health.

## Data Availability

The datasets generated during and/or analyzed during the current study will be available from the principal investigator (H. Irene Su) on reasonable request.

## Author Contributions

Dayo, Reyes, Flores Ortega, Kaiser, Aarons, McMenamin, Su, Romero – Conceptualization and Methodology

Dayo, Turcotte, Reyes, Flores Ortega, McMenamin, Su, Romero, – Investigation and Data curation

Dayo, Flores Ortega, Kaiser, Su, Romero, - Formal analysis

Dayo, McMenamin, Su, Romero - Project administration

McMenamin, Su - Resources

Dayo, Kaiser, McMenamin, Su, Romero – Supervision

Dayo, Turcotte, Reyes, Romero - Writing – original draft

All Authors - Writing – review & editing

## Appendix

Sample of health insurance educational guide highlighting key health insurance literacy terms and considerations when choosing among the five health plan options for in vitro fertilization coverage given to participants randomized to the Intervention arm. Participants randomized to the Usual Care arm were given the last page (Appendix 1).

---

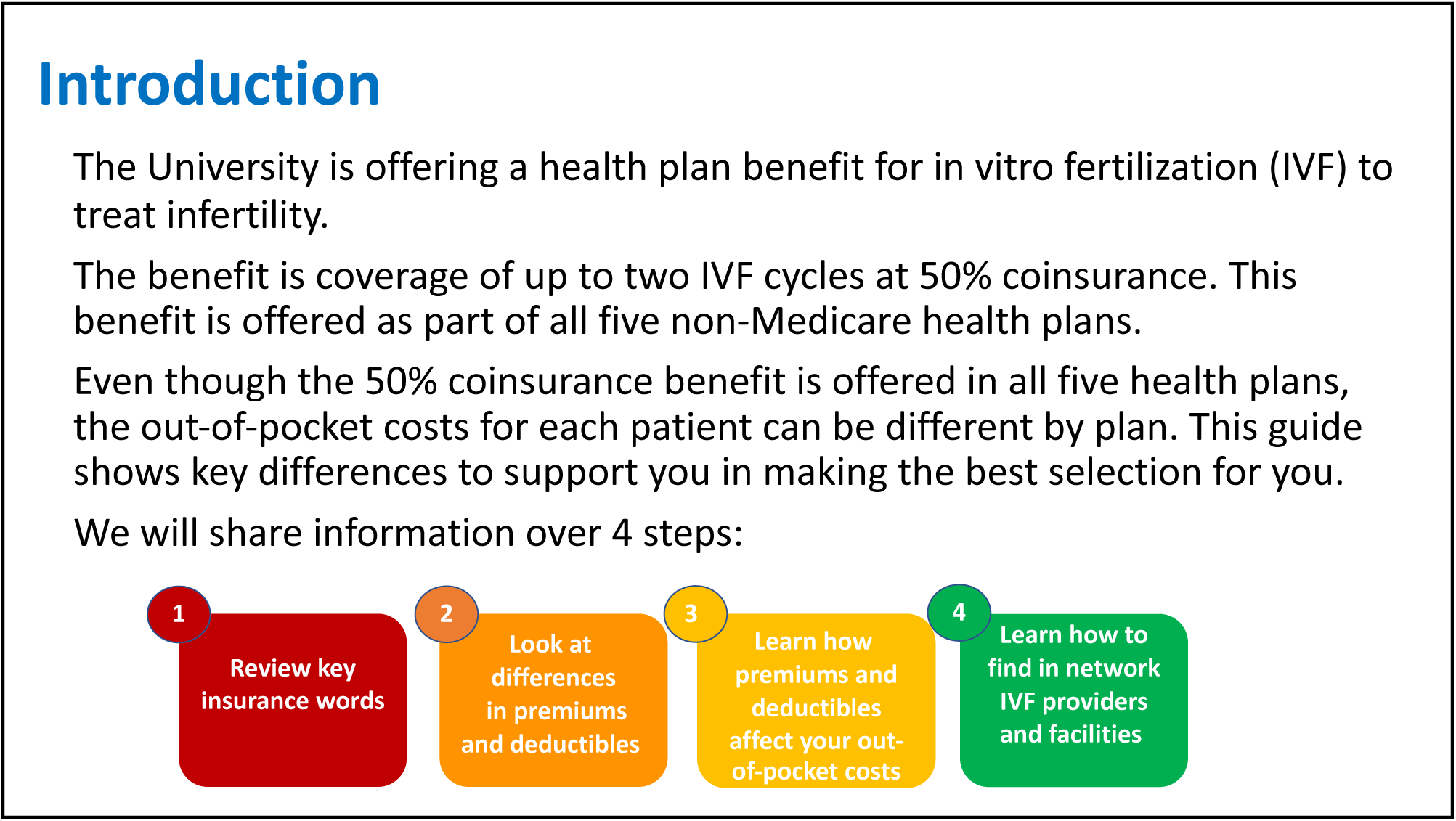

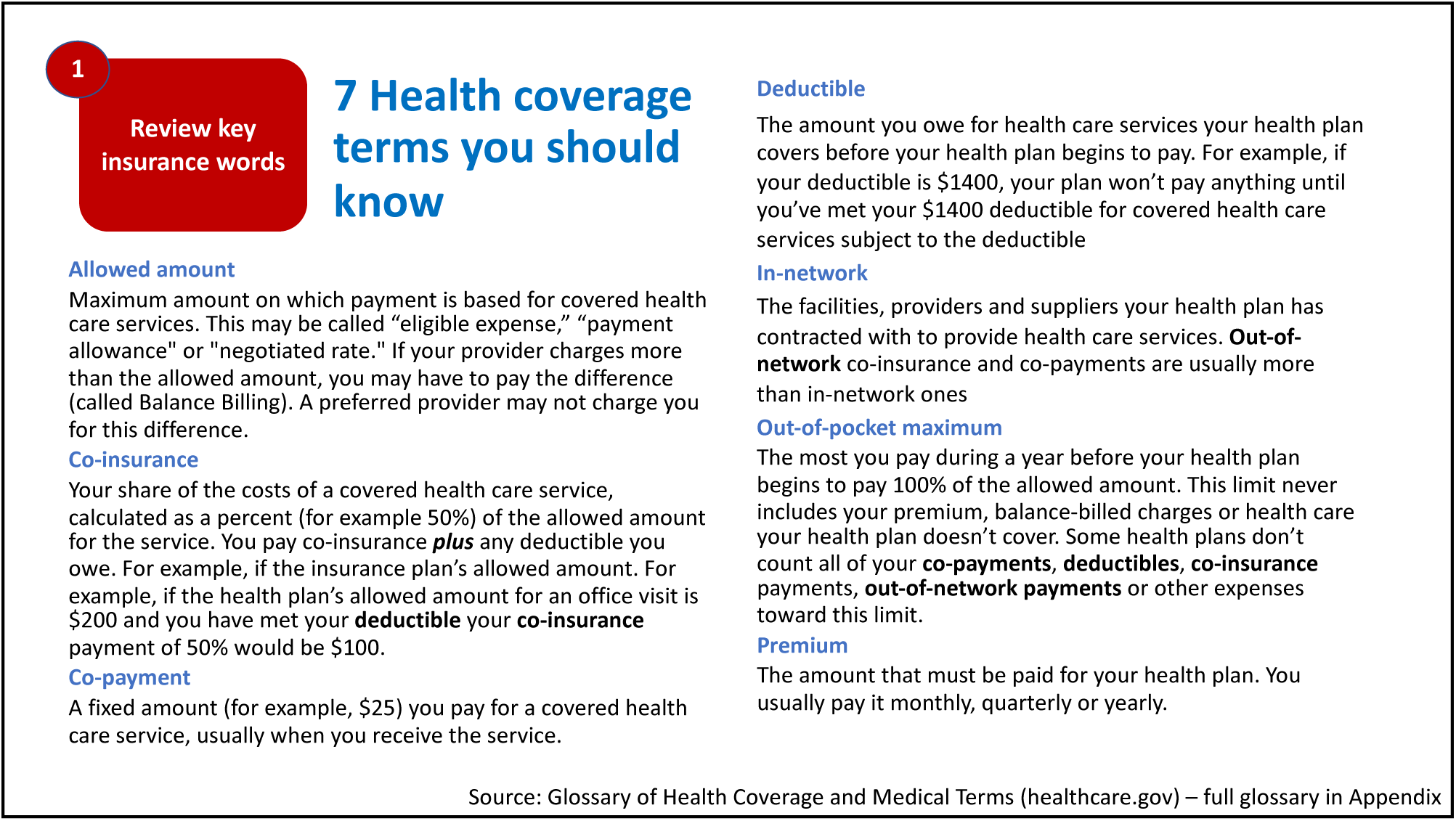

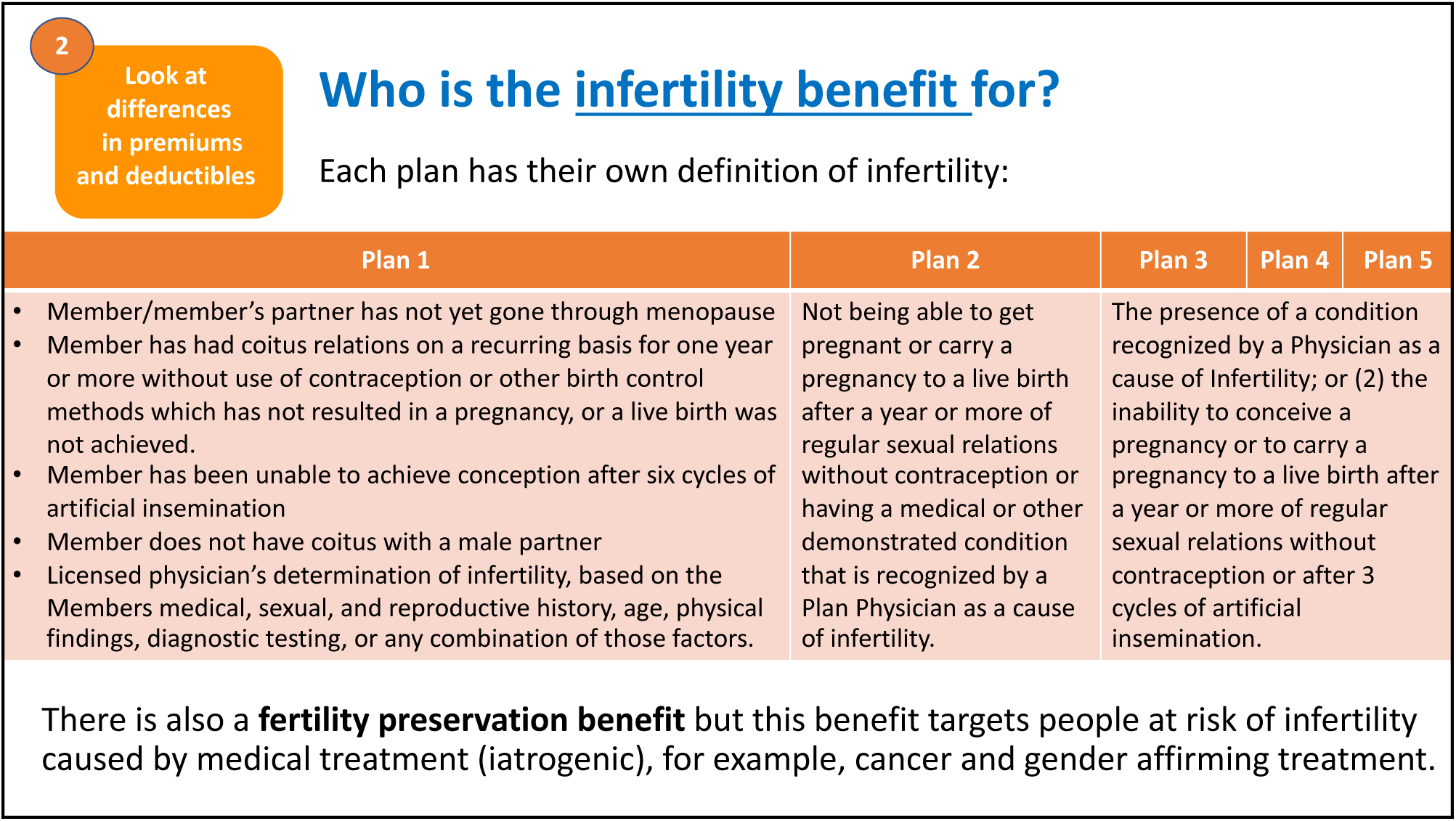

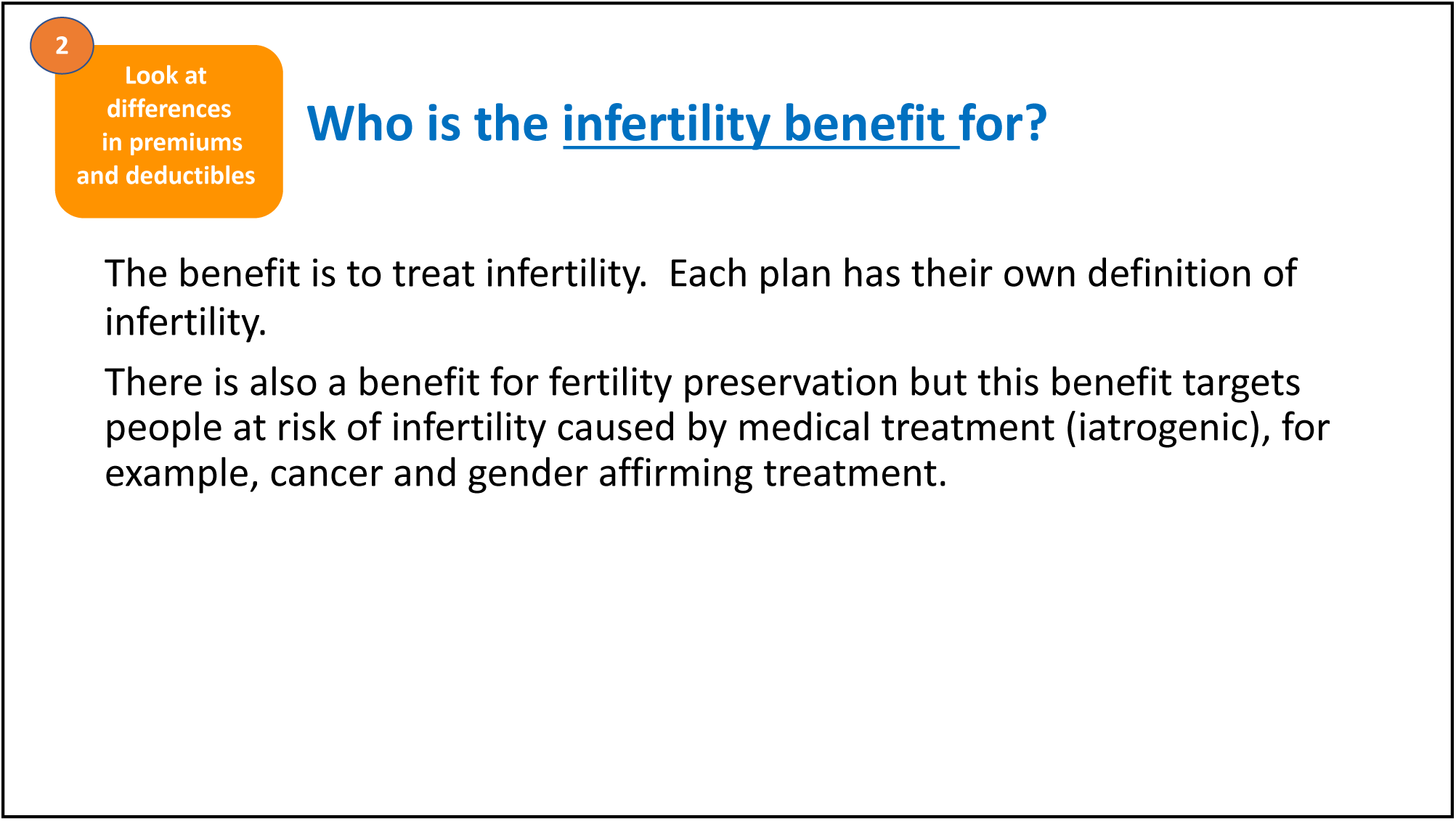

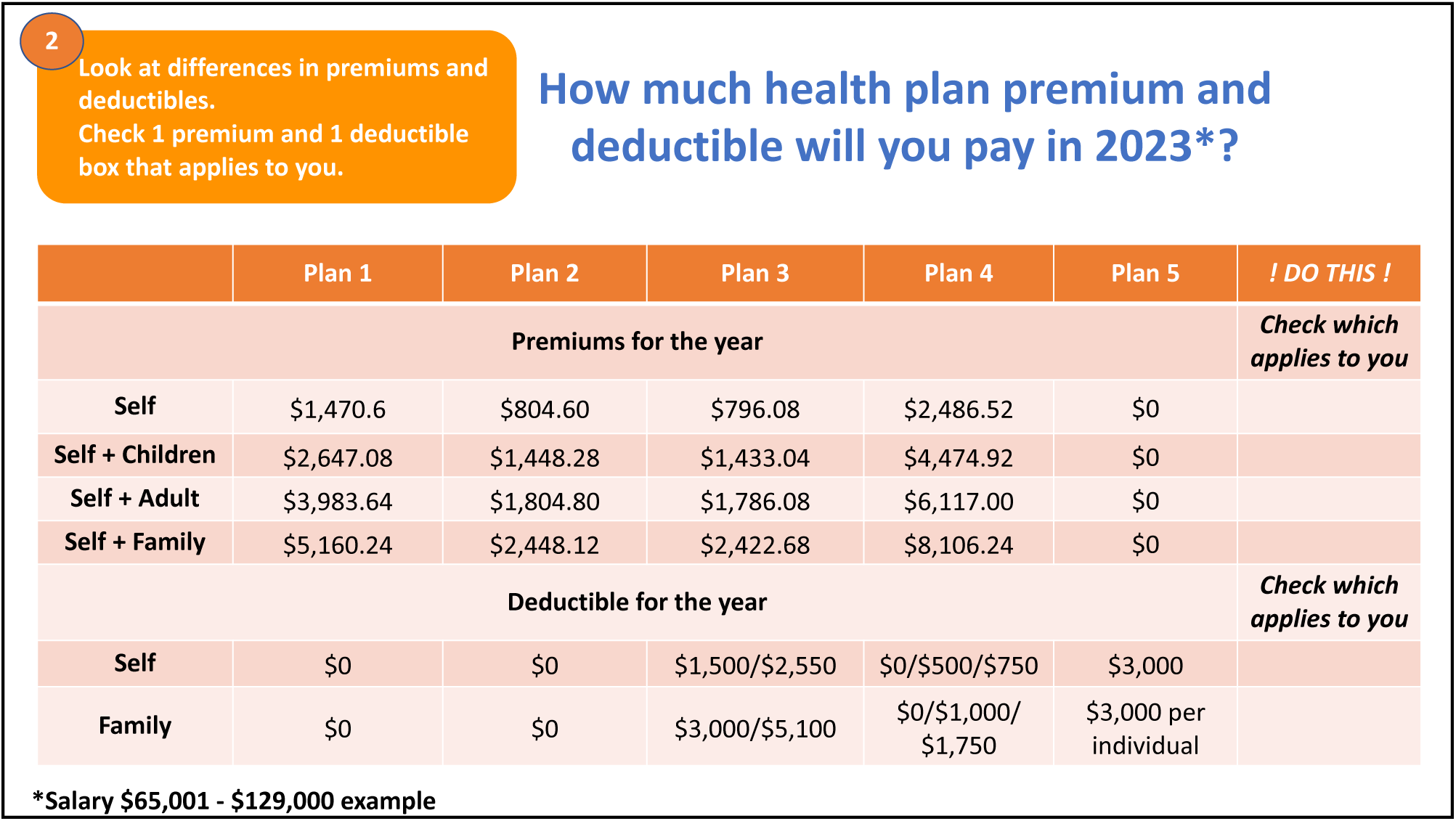

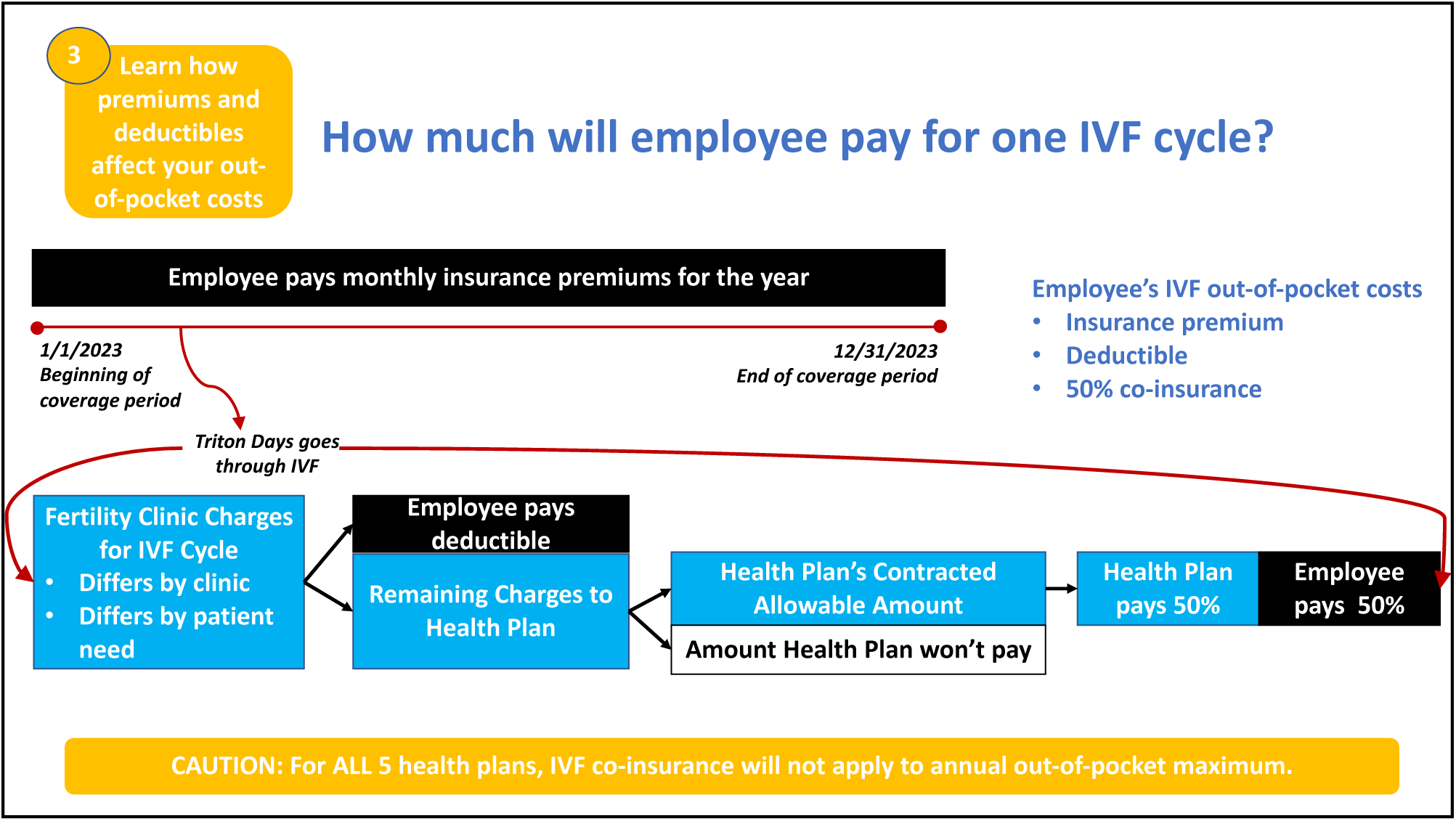

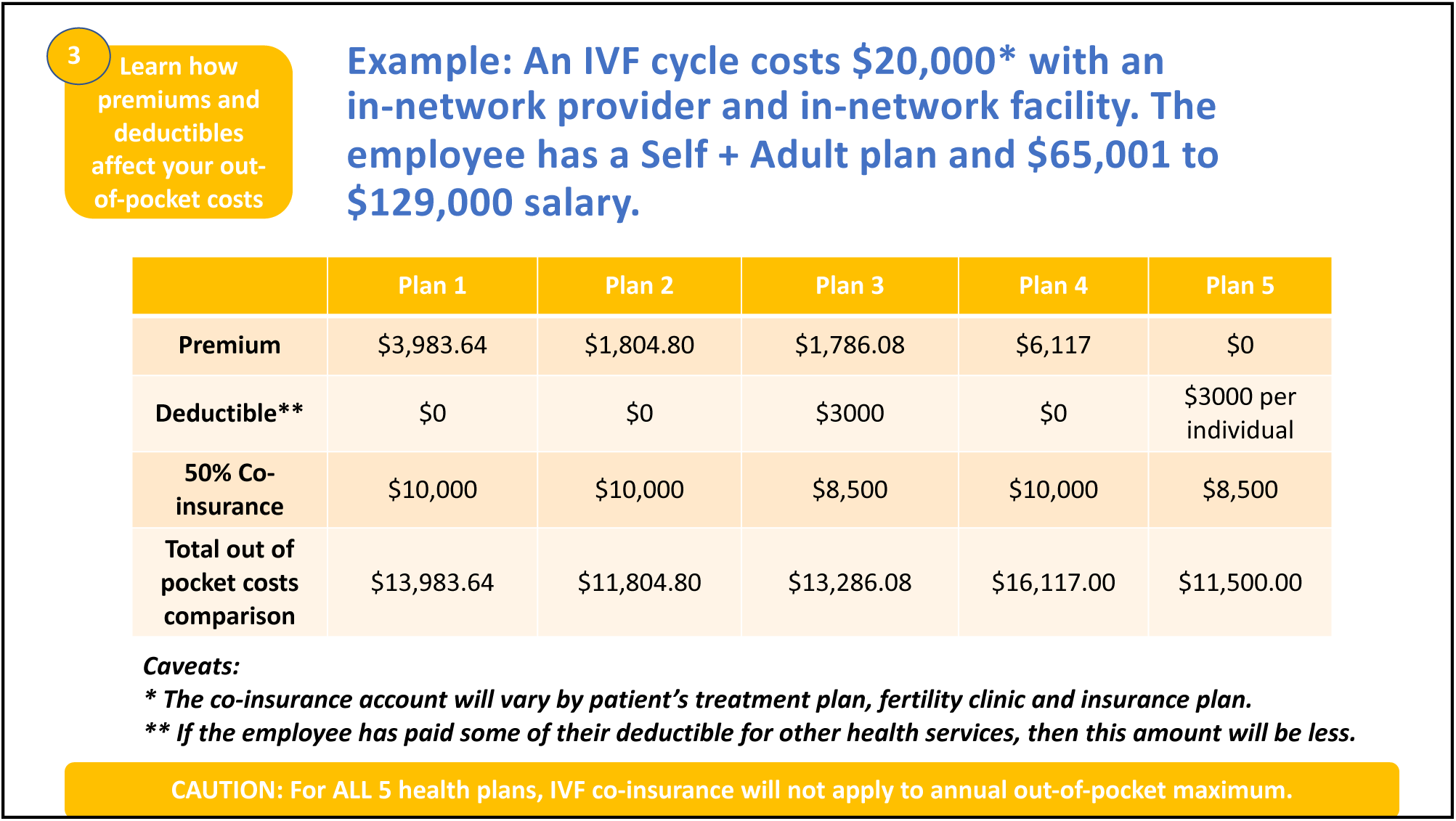

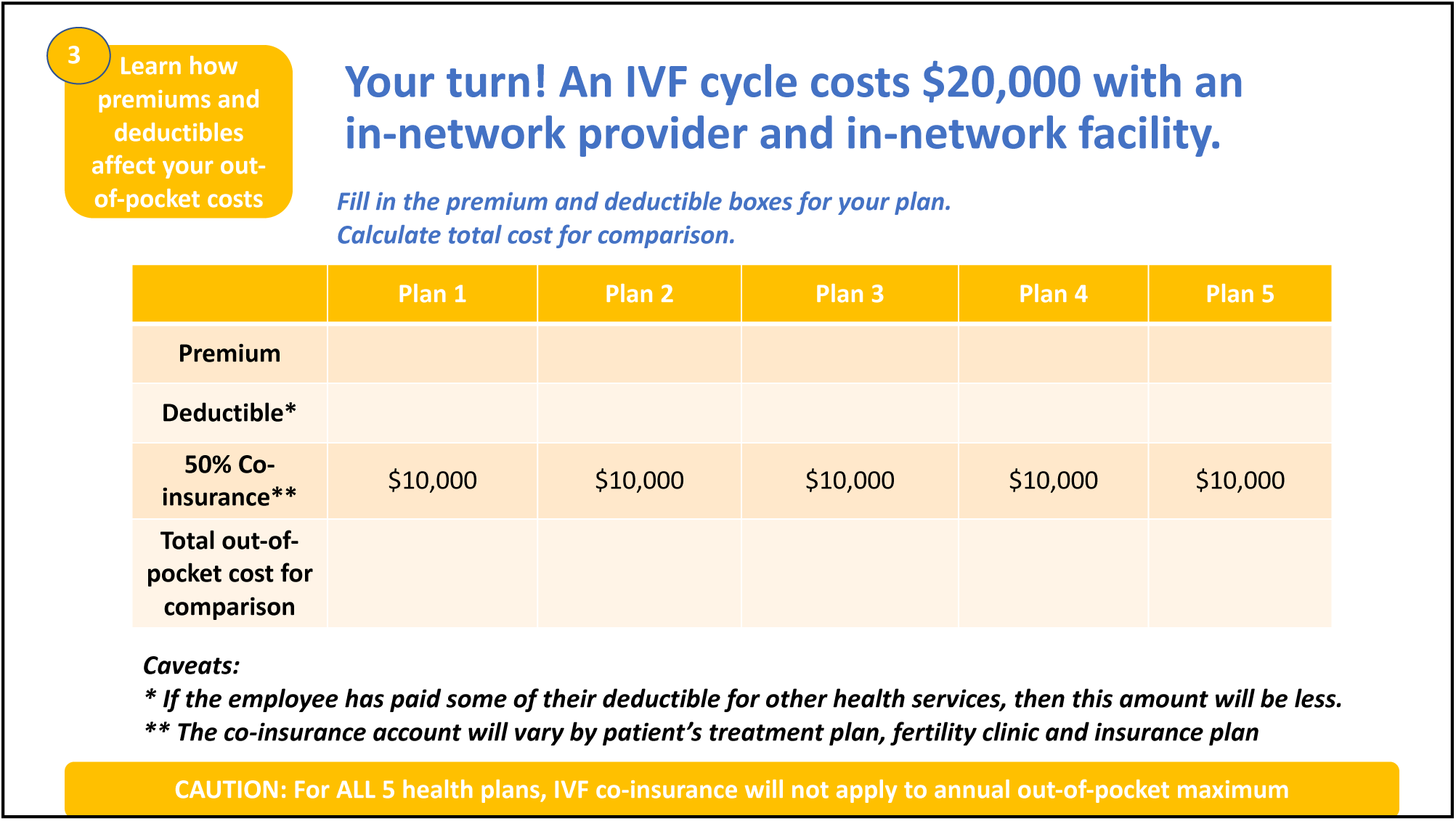

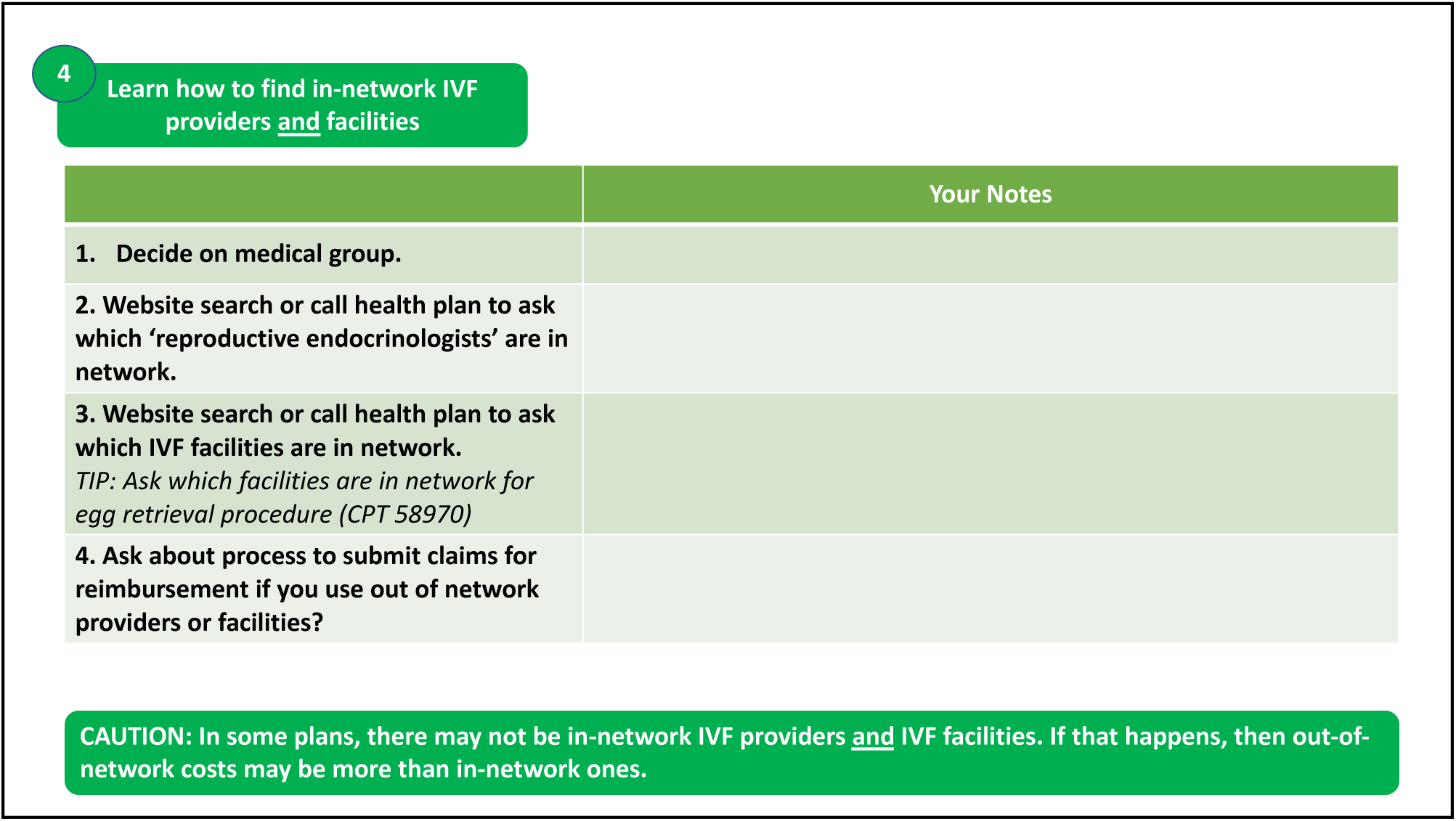

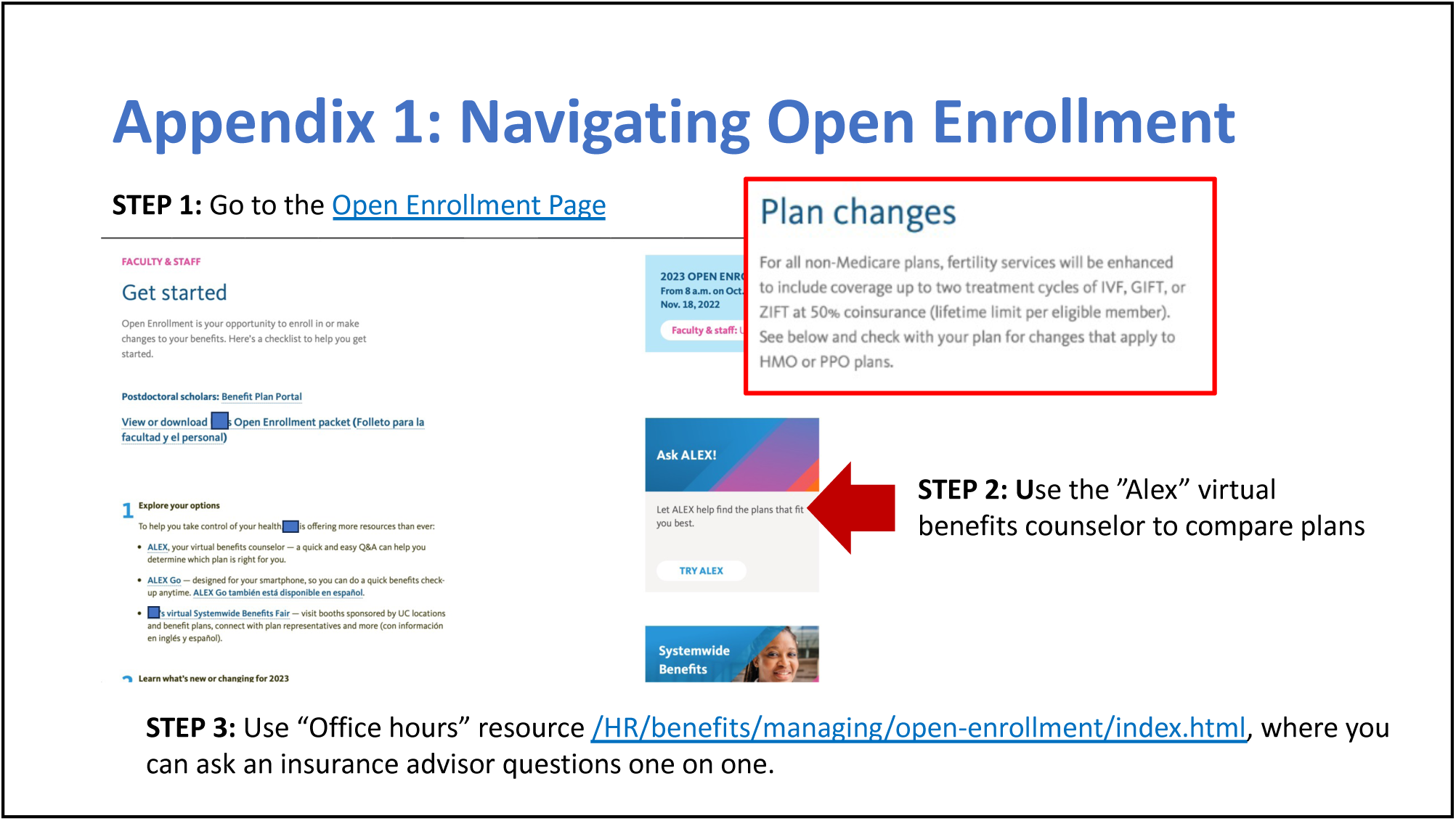

